# A potential impact of physical distancing on physical and mental health. A rapid narrative umbrella review of meta-analyses on the link between social isolation and health

**DOI:** 10.1101/2020.10.06.20207571

**Authors:** Nexhmedin Morina, Ahlke Kip, Thole H. Hoppen, Stefan Priebe, Thomas Meyer

## Abstract

**Background:** The imperative for physical distancing during the coronavirus disease 2019 (COVID-19) pandemic may deteriorate physical and mental health. We aimed at summarizing the strength of evidence in the published literature on the association of social isolation and loneliness with physical and mental health.

**Methods:** We conducted a systematic search in April 2020 to identify meta-analyses using the Medline, PsycINFO, and Web of Science databases. The search strategy included terms of social isolation, loneliness, living alone, and meta-analysis. Eligible meta-analyses needed to report any sort of association between an indicator of social isolation and any physical or mental health outcome. The findings were summarized in a narrative synthesis.

**Results:** Twenty-five meta-analyses met our criteria, of which 10 focused on physical health and 15 on mental health outcomes. A total of more than 3 million individuals had participated in the 692 primary studies. The results suggest that social isolation is associated with chronic physical symptoms, frailty, coronary heart disease, malnutrition, hospital readmission, reduced vaccine uptake, early mortality, depression, social anxiety, psychosis, cognitive impairment in later life, and suicidal ideation.

**Conclusions:** The existing evidence clearly indicates that social isolation is associated with a range of poor physical and mental health outcomes. A potential negative impact on these outcomes needs to be considered in future decisions on physical distancing measures.

**Strengths and limitations of this study:**

- This rapid umbrella review focuses on a timely and societally relevant issue.
- The systematic literature search was conducted in three major databases from inception up to April 2020 warranting an extensive and up-to-date overview on relevant meta-analyses in the field.
- Quality of included meta-analyses was rated with a standardized measure.
- Different indicators of social isolation were included.
- The utilized method did not allow for a quantitative comparison of associations with health outcomes.

## Background

The coronavirus 2019 (COVID-19) pandemic poses a global public health threat. In order to slow the spread of the virus by reducing contact rates, governments around the world have taken unprecedented political decisions that have transformed societies. The exact form and extent of these measures have varied, but they always include some type of physical distancing (mostly referred to as social distancing) making it impossible for people to maintain their normal social life.

In many countries, the restrictions have already been in place for several months. Depending on the further course of the pandemic with potential new waves, restrictions might continue for longer periods of time or be re-imposed after periods of loosening or abandoning them. When deciding about imposing, continuing or relaxing measures of physical distancing, governments have to consider and balance different risks. Whilst physical distancing is likely to reduce the risk of spreading the virus, it might generate other risks. These include potential damages to the economy and also possible negative consequences for the health of the population. For a balanced decision on further physical distancing measures, evidence is required on whether the measures are likely to impact on a range of health outcomes.

A recent general population survey revealed that physical distancing can increase social isolation and loneliness.^1^ This may happen when people are prevented from travelling, physical meetings with significant others, and in some cases even from leaving their home other than for essential activities. Of note, some individuals can be physically isolated and not feel lonely and others can feel lonely even if they are not isolated. Furthermore, many individuals are able to remain socially connected by means of remote communication while physically isolated. Accordingly, we should not assume that physical distancing inevitably leads to social isolation and loneliness. However, physical distancing is likely to have a disproportionate effect on those most vulnerable, in particular older adults, individuals in need of intensive physical or mental health care, and individuals with limited access to technology who lack the means of engaging in creative forms of contact with loved ones. Older patients, for example, may lose access to important parts of their usual routine (e.g., day care programs or informal gatherings with significant others). Similarly, caregivers residing with patients need also to physically isolate themselves due to the ramifications of quarantines.

Social isolation is a broad term without a consistent definition in the literature. Three indicators of social isolation (also referred to as social connections) are commonly used in research: few social network ties, living alone, and loneliness.^2-4^ Social network ties is a behavioral measure that can – at least in theory – be objectively quantified. Living alone describes a basic characteristic of an individual’s social situation which can be associated with reduced social relationships, but is not necessarily so.^5^ Loneliness, on the other hand, is an individual’s subjective assessment of the quality and quantity of their social relationships, reflecting a belief that they have too few or too poor relationships, or both. Accordingly, social network ties and living alone represent structural indicators, whereas loneliness represents a quality measure of social connections.^4,5^

Although these three indicators capture distinct aspects of social isolation, they commonly overlap and are associated with each other. Literature suggests that many individuals are socially isolated or lonely or both and that social isolation and loneliness may occur unequally across age groups. For example, Hawkley and colleagues^6^ reported that loneliness decreased with age through the early 70s and then increased again. Several studies indicate that at least a fifth of adults report frequent loneliness,^7,8^ and that more than 40 percent of adults aged 60 and older report feeling lonely.^9^

The extent to which individuals are socially isolated can have a profound impact on both physical and psychological well-being.^5^ Social isolation is thought to influence health through behavioral and biological pathways.^10^ Several studies demonstrate that social isolation is associated with health-relevant behaviors, such as lack of physical activity, poorer sleep, obsessive behavior, as well as neuroendocrine dysregulation,^10^ chronic allostatic load,^11^ high blood pressure and poor immune functioning.^5,12,13^ Furthermore, the magnitude of the effect of social isolation on mortality may be equivalent to or exceed the impacts of deleterious behaviors such as excessive drinking or obesity.^3^

Physical distancing may increase social isolation therefore have a negative impact on physical and mental health. For weighing up this potential impact in policy decisions, the existing evidence needs to be considered. Against this background, we conducted a systematic umbrella review to synthesize the evidence on the association between social isolation and physical and mental health outcomes. As recommended by the World Health Organization (WHO), we explored relevant meta-analyses by means of a rapid review of evidence.^14^

## Methods

The aims and methods of this umbrella review were registered with the PROSPERO database (http://www.crd.york.ac.uk/prospero). To select relevant meta-analyses on the association between social isolation and physical or mental health outcomes we conducted a systematic search on 6^th^ April 2020 using the databases Medline, PsycINFO, and Web of Science. We conducted multi-field searches (in titles, abstracts, and key concepts) using the following terms: social isolation, loneliness, living alone, and meta-analy*, which we combined using the Boolean operators “or” plus “and”. The full search string for Medline and PsycINFO was “((TI Loneliness OR AB loneliness OR SU Loneliness) OR (TI social isolation OR AB social isolation OR SU social isolation) OR (TI living alone OR AB living alone OR SU living alone)) AND (TI meta-analy* OR AB meta-analy* OR SU meta-analy*)”. Relevant outcomes included any sort of physical or mental health outcome. We applied no restrictions on age of participants, applied research designs (i.e., cross-sectional, longitudinal), or publication language. Furthermore, we did not apply any limits. We first inspected the title and abstract of all hits and then read full texts of the hits that seemed to meet the aforementioned inclusion criteria. The Preferred Reporting Items for Systematic Reviews and Meta-analyses reporting standards were followed to document the process of systematic review selection. ^15^

### Coding of trial characteristics

Systematic reviews with a quantitative synthesis of trial results (meta-analysis) were retained. Two reviewers (NM & THH) coded and extracted from each meta-analysis several objectively verifiable characteristics: Authors and year of publication, inclusion criteria, number of included primary studies, number of participants and their composition by age and health conditions, study design, type of social connection (social network ties/living alone/loneliness) evaluated, clinical outcome, length of follow-up, number of databases searched, and search areas. Furthermore, we extracted the main findings on the association between social network ties/living alone/loneliness and health outcomes (correlation values, odds ratios, or hazard ratios, and the corresponding 95% confidence intervals). With respect to the 95% confidence intervals, both values greater than one (or both values less than one) represent a significant increase (or decrease) as a function of social isolation.

### Quality Assessment

The quality of included systematic meta-analyses was independently assessed by two reviewers (AK & TM) using A Measurement Tool to Assess Systematic Reviews – 2 (AMSTAR-2).^16^ Following the tool’s guidelines, the raters assigned one of four global quality ratings (i.e., high, moderate, low, or critically low) after consideration of 16 potential critical and non-critical weaknesses. Items addressing the following criteria were considered as critical: Clear research question including definitions of population, intervention, control group, and outcomes (PICO), adequacy of the literature search, and adequate assessment and/or consideration of risk of bias in the primary studies. Typically, high and moderate ratings reflect the presence of one or more non-critical weakness, while low and critically low ratings indicate one or more critical weaknesses. Any discrepancies among the independent raters were discussed until consensus was reached.

## Results

### Selection and characteristics of included studies

Figure 1 displays a PRISMA^15^ flow diagram of the publication selection process. After reading 530 abstracts, 89 full text publications were reviewed. The final review resulted in 25 meta-analyses. Relevant characteristics of these meta-analyses are summarized in Table 1.

**Table 1:**
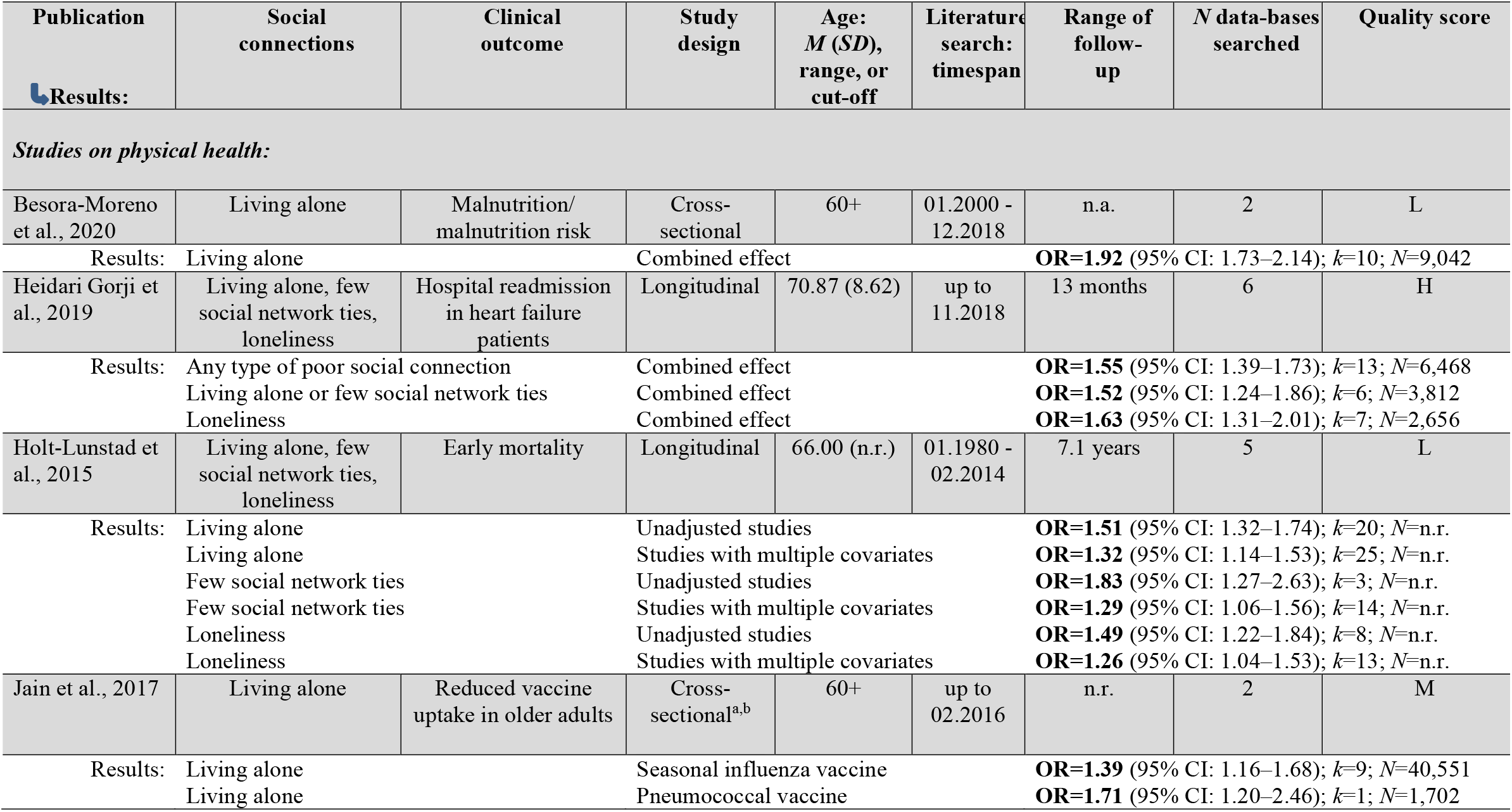
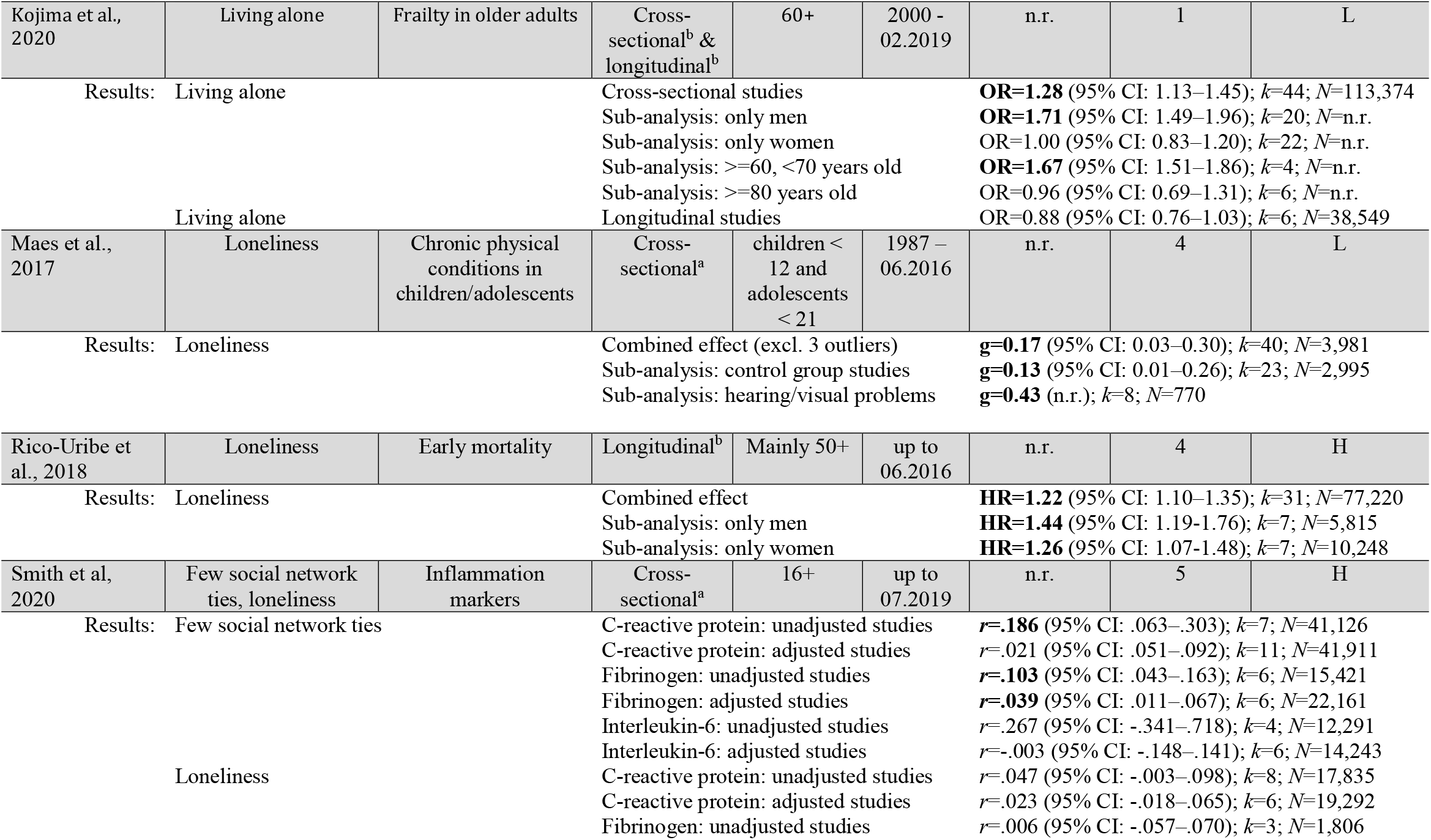
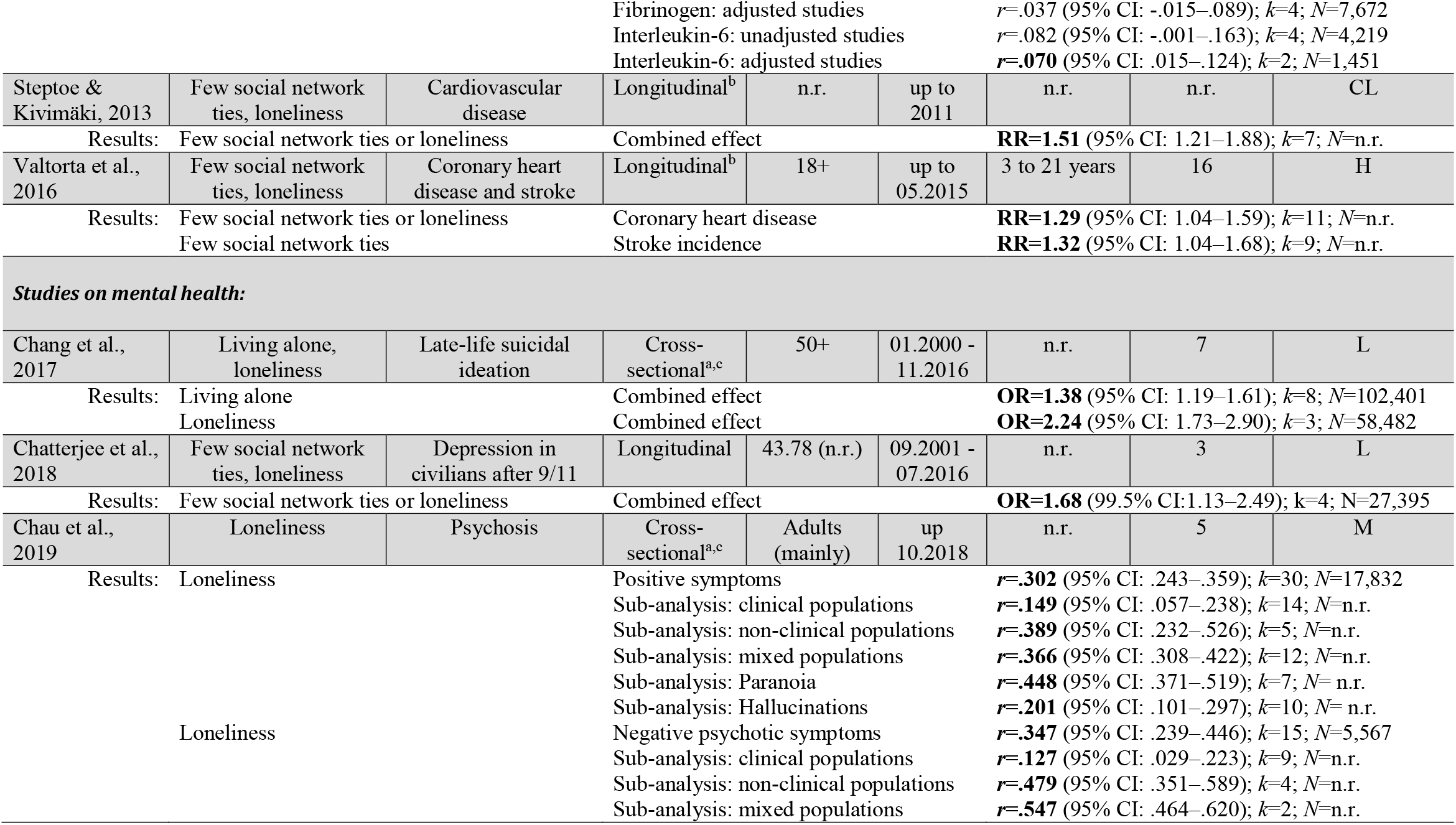
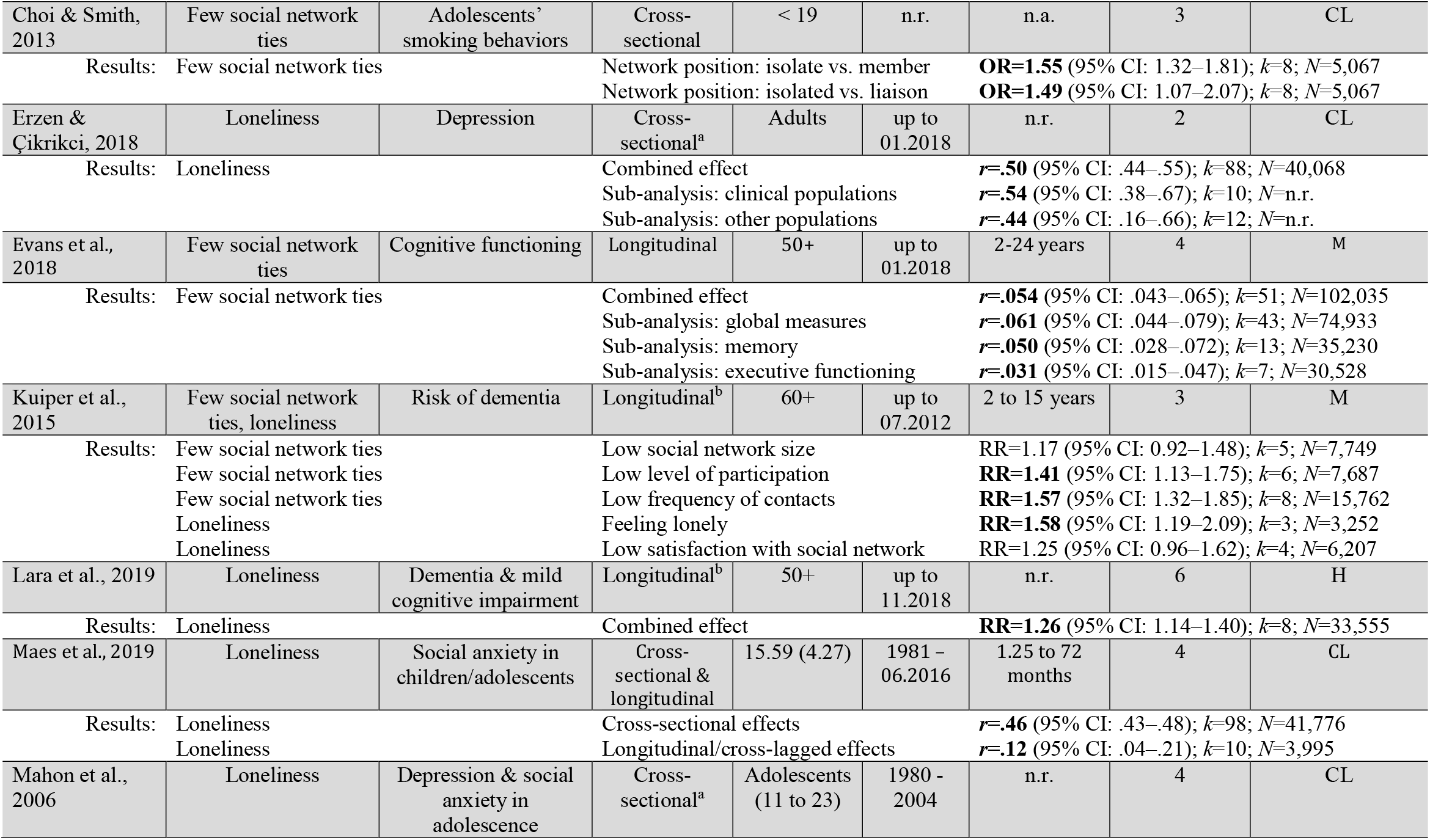
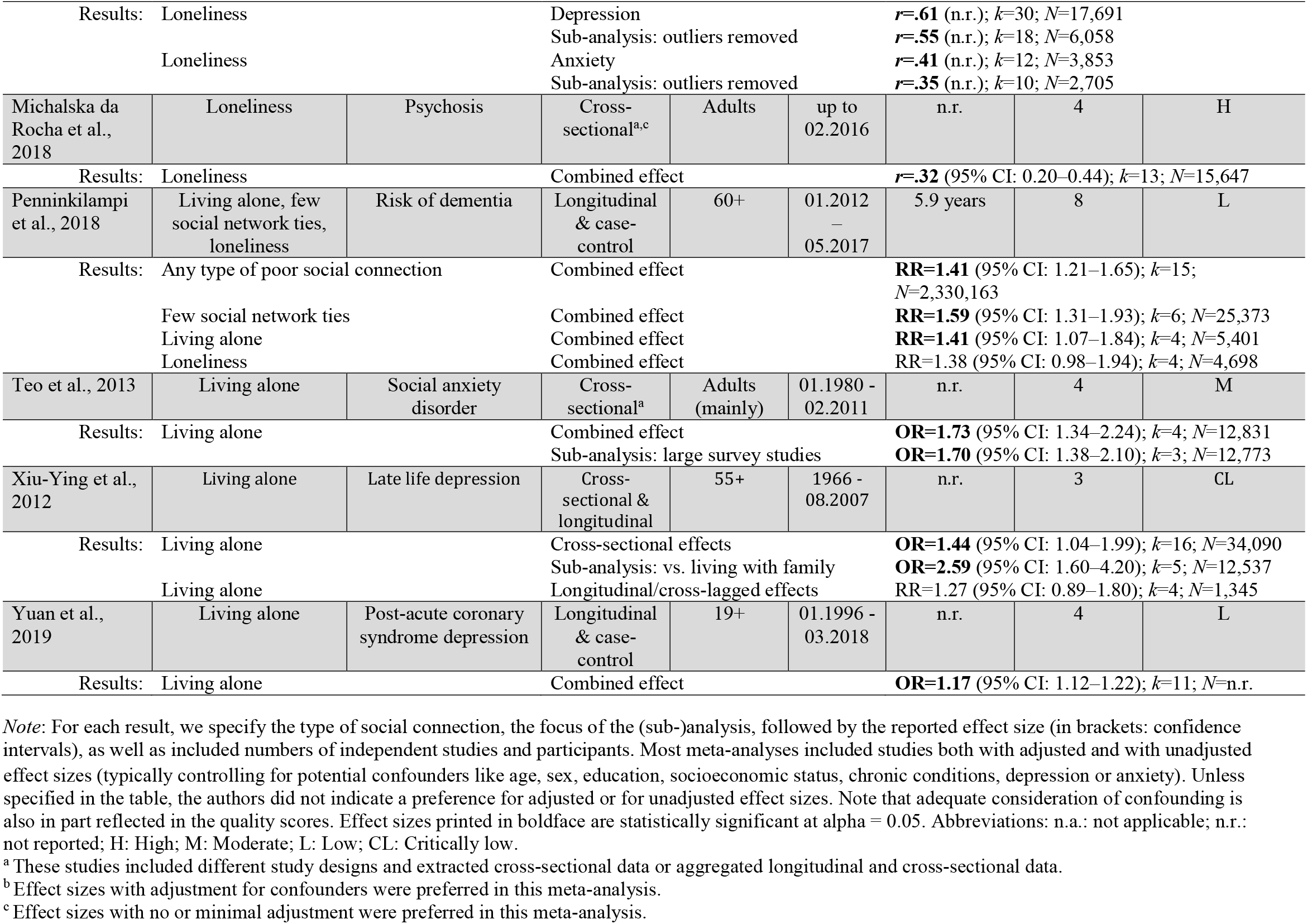
Overview of the included meta-analyses

**Fig. 1.**
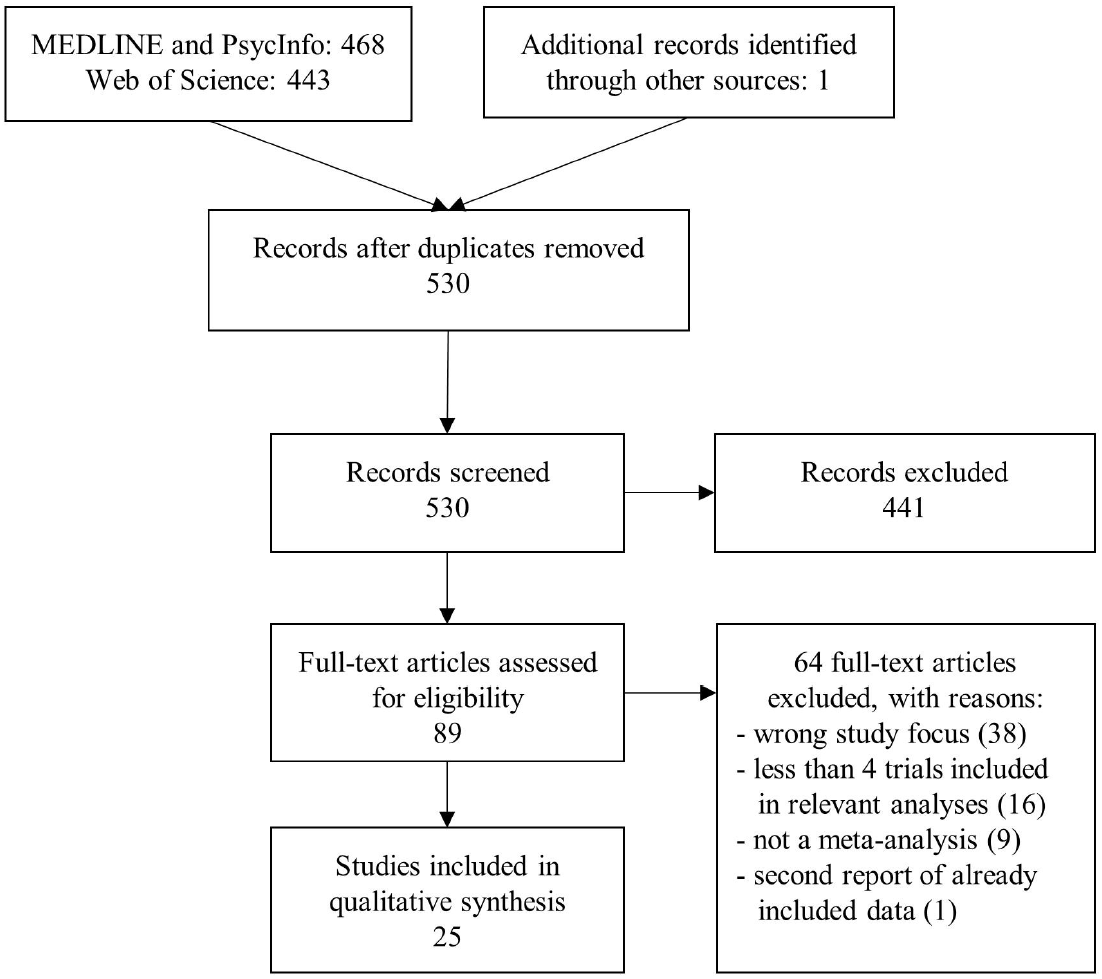
Flow diagram of study selection process.

All publications were journal articles in English. Ten meta-analyses reported associations of few social network ties, living alone, and loneliness with physical health outcomes, and 15 with mental health outcomes. Different indicators of social isolation were measured in the included studies. We considered as structural indicators of social isolation *social network ties* defined as an objectively quantifiable variable of one’s social contacts irrespective of its perceived quality and *living alone* as an objective characteristic of the living situation. Furthermore, we defined *loneliness* as a quality indictor representing the subjective emotional appraisal of the extent and quality of social relationships.^5^ The meta-analyses differed with respect to whether they kept these three measures of social isolation separate of whether they combined them (see Tab. 1).

A total of 276 primary studies were included in the 10 meta-analyses on physical health. The reported results in Table 1 were based on sample sizes ranging from 1,451^17^ to 113,374^18^ participants, with three meta-analyses not reporting on the sample size. Five meta-analyses were based on longitudinal studies only, one on cross-sectional studies only, and the remaining four on a pooled combination of both cross-sectional and longitudinal studies. Furthermore, social network ties and living alone were examined in 5 meta-analyses on physical health, respectively. Loneliness, on the other hand, was examined in 7 meta-analyses on mental health. Only one of these studies was conducted with children and adolescents.^19^ The meta-analyses based on cross-sectional studies revealed a significant association between social isolation and the following health problems: chronic physical complaints in children and adolescents,^19^ coronary heart disease and stroke,^20^ and frailty in older male (but not female) adults.^18^ Additionally, social isolation was associated with malnutrition^21^ and vaccine uptake amongst older adults.^22^ One meta-analysis^17^ reported mostly non-significant results on a positive association between social isolation and inflammation (acute-phase C-reactive protein and fibrinogen). The meta-analyses conducted with longitudinal studies indicate that social isolation is associated with increased risk of cardiovascular disease,^23^ early mortality,^3,24^ and hospital readmission in heart failure patients.^25^

The 15 meta-analyses on mental health were based on a total of 416 primary studies. The reported results are based on sample sizes ranging from 1,345^26^ to 2,330,163^27^ participants, with one meta-analysis failing to report on the sample size. Four of the 15 meta-analyses provided longitudinal data only, one provided cross-sectional data only, and the remaining ten meta-analyses reported on both cross-sectional and longitudinal studies. In addition, social network ties, living alone, and loneliness were examined in 5, 5, and 10 meta-analyses on mental health, respectively. Three meta-analyses focused on studies with children and adolescents.^28-30^ The included meta-analyses based on cross-sectional designs reported a significant positive association between social isolation and late-life suicidal ideation,^31^ depression in adults,^32^ late life depression,^26^ psychosis,^28,33,34^ smoking behavior in adolescents,^29^ depression and social anxiety in childhood and adolescence,^28,30^ and social anxiety disorder in adults.^35^ The meta-analyses based on longitudinal studies suggest that social isolation is associated with higher risk of depression in adults,^36^ post-acute coronary syndrome depression,^37^ and dementia and cognitive impairment in later life.^27,38-40^ See Table 1 for detailed information.

### Study quality

The Intraclass Correlation Coefficient (ICC) of the global quality ratings among the two raters was .83, 95% CI = .62 – .93, indicating good inter-rater reliability. Study quality was very heterogeneous among meta-analyses both on physical and mental health (see Tab. 1). With respect to the meta-analyses on physical health, the global rating was high in 40%, medium in 10%, low in 40%, critically low in 10% of the meta-analyses. In the 15 meta-analyses on mental health, study quality was rated as high in 13%, medium in 27%, low in 27%, and critically low in 33% of the meta-analyses. Among the AMSTAR-2 criteria, inadequate assessment of risk of bias and/or lack of consideration of risk of bias represented the most frequent critical weaknesses of included meta-analyses.

## Discussion

The review clearly demonstrates that social isolation is associated with poorer health. This applies to a range of physical and mental health outcomes and has been found in different populations and contexts. The evidence based on both cross-sectional and longitudinal data is substantial for physical health outcome and even more extensive for mental health outcomes. More specifically, social isolation is linked with chronic physical symptoms, frailty, coronary heart disease, stroke, early mortality, malnutrition, hospital readmission in heart failure patients, and vaccine uptake. With respect to mental health, social isolation is linked with depression in young and adult populations, social anxiety, psychosis, dementia and cognitive impairment in later life, and late-life suicidal ideation.

### Strengths and limitations

This is, to our knowledge, the first review to synthesize the existing evidence that has been reported in meta-analyses on the link between social isolation and physical and mental health outcomes. The findings reflect a reasonable number of meta-analyses which in total included 692 studies. Thus, the overall conclusions of this umbrella review are based on an extensive body of empirical evidence.

However, the review also has several limitations. Firstly, we considered different indicators of social isolation, and our method did not allow us to identify whether one indicator is more relevant than another. Secondly, half of the included meta-analyses for both physical and mental health outcomes had an overall quality rated on AMSTAR-2 as low or critically low, with inadequate consideration of risk of bias being the most frequent critical flaw. Thirdly, the quality of the primary research studies that went into the included meta-analyses also varied and their different methodological shortcomings cannot be adequately considered in this review. Fourthly, the results on the association between living alone and health outcomes need to be interpreted with caution. As reported above, living alone is not necessarily indicative of feeling lonely.^2^ Finally, the review included a wide range of health outcomes and did not quantify the strength of the associations for different outcomes.

### Implications

The review leaves little doubt that social isolation is linked with poorer physical and mental health. The findings are strengthened by the fact that several meta-analyses were conducted with longitudinal studies. In particular, longitudinal data indicate that social isolation is associated with increased risk of several physical and mental health outcomes, cardiovascular disease, hospital readmission in heart failure patients, early mortality, cognitive impairment, and depression.^3,23-25,27,36-40^ However, the findings are all based on observational studies and do not provide evidence on the causal direction of the association. Poor physical and mental health can lead to social isolation, and social isolation can lead to poorer health. For establishing a causal relationship and examining the strength of the predictive relationship of social isolation and loneliness with health outcomes experimental studies are required, which were not the subject of this review.^4,41^ Experimental research with animals, however, suggests that social isolation increases mortality.^42^ Furthermore, experimental studies with humans indicate that randomly inducing loneliness or exclusion leads to different health relevant physiological responses than being randomly assigned to a support condition.^42^ For most of the considered outcomes in this review, a causal effect of social isolation is plausible and likely to explain at least part of the identified associations. The casual direction is definite in case of the greater risk of isolated people to die early.^3^ For an explanation of the damaging effect of social isolation on health outcomes, one may refer to different theoretical models. Theorists from different perspectives have postulated that the impact of social isolation on health is mediated by impairments in social capital,^43^ social control,^44^ social identification,^45^ and social support.^46^

Furthermore, some evidence from randomized controlled trials, however, suggests that expanding the social connections of individuals, e.g., through befriending programs, may indeed improve different health outcomes.^47^ Altogether, the literature on interventions to reduce loneliness and social isolation indicates that a policy focus on social connection is a cost-effective strategy for enhancing health at the population level due to the potential pay-offs in health care costs that would otherwise occur. Existing volunteer friendly visiting programs or psychosocial group interventions^48^ may need to be redesigned to the point that they can be readily implemented in in accordance with existing rules of physical distancing. Creative programs and interventions to foster social connections, including technology-based social networking programs, are needed.^49^ Furthermore, existing policies should ensure that populations at greater risk, such as the poor and the elderly, receive most support.^1^

All the included studies assessed social isolation as it occurs in a normal societal context. Physical distancing as part of measures to limit the spread of COVID-19 is different from the situations considered in the research synthesized in this review. Firstly, for the vast majority of the population, the required physical distancing leads to a much more pronounced social isolation than what they have experienced before. Secondly, physical distancing is externally imposed and not due to individual life style decisions, lack of material means, poor social skills or other barriers to socialize. And thirdly, physical distancing is requested from people in an overall context of uncertainty that comes with further stressors, health risks, and often a reduced accessibility of health care.

It is important to note that physical distancing is a broad umbrella term that incorporates a wide range of potential measures, with highly divergent implications for social routines. It can include a full lock down and curfew, specific guidelines for meetings and gatherings of people, physical distancing in public, and a recommended or mandatory wearing of face masks. The type, degree, and duration of physical distancing measures have been variable across countries and will affect how isolated different groups in the population become.

One can only speculate as to whether and, if so, to what extent the increased social isolation resulting from physical distancing measures in the current situation will have an even greater impact on health outcomes than has been suggested in this review. Arguably, an even greater impact can be expected for certain risk groups, such as older people who are more threatened by COVID-19 and socially disadvantaged groups (e.g., individuals in need of mental or physical health care or individuals with low income) who often face even more economic adversity than before the pandemic. Further research is required to identify which populations are at particular risk to suffer health problems as a result of physical distancing and to explore whether the resulting social isolation may – at least to some extent and in some people – be compensated through positive effects of the pandemic, such as strengthened local communities and increased options for online social activities.^47,50^

## Conclusions

In governmental decisions about future physical distancing measures, a potential negative impact of the resulting physical isolation on the health of the population needs to be considered. The existing literature suggests that social isolation and loneliness may affect both physical and mental health outcomes and include an excess mortality. However, the potential impact of physical distancing on social isolation and loneliness and ultimately on physical and mental health outcomes need to be thoroughly examined. In addition, the existing knowledge on the association between social connection and physical and mental health should be considered in clinical practice. Finally, more experimental research is needed to increase our understanding of the causal relationship between social connection and physical and psychological well-being.

## Data Availability

All data generated or analysed during this study are included in this article.

## Acknowledgements

Not applicable.

## Report about dual (co-)authorship

No authors co-authored any of the systematic reviews and meta-analyses included in our overview.

## Authors’ Contributions

NM had full access to all of the data in the study and takes responsibility for the integrity of the data and the accuracy of the data analysis. NM designed the search strategy with input from AK and TM. NM and AK carried out the literature searches and screening. NM, THH, and TM carried out the data extraction. AK and TM assessed the quality of the included meta-analyses. NM and SP wrote the first draft of the manuscript and all authors contributed to and have approved the final manuscript.

## Funding/Support

No funding received.

## Conflict of Interest Disclosures

Not applicable.

## Availability of data and materials

All data generated or analysed during this study are included in this published article and supplementary material.

Ethics approval and consent to participate: This study did not require ethical approval.

## Consent for publication

Not applicable.

## Patient and Public Involvement statement

Not applicable.

## Notes

### Competing Interest Statement

The authors have declared no competing interest.

### Author Declarations

This umbrella review of published meta-analyses did not require ethical approval.

